# An individualized digital twin of a patient for transdermal fentanyl therapy for chronic pain management

**DOI:** 10.1101/2022.07.25.22277536

**Authors:** Flora Bahrami, Rene Michel Rossi, Katelijne De Nys, Thijs Defraeye

## Abstract

Fentanyl transdermal therapy is a suitable treatment for moderate-to-severe cancer-related pain. Patients show inter-individual drug uptake behavior, which leads to a different response to the therapy. This study aims to determine the effect of different physiological features of the patients on the pain relief achieved with the therapy. Therefore, a set of virtual patients was developed by using Markov Chain Monte Carlo (MCMC), based on actual patient data. The members of this virtual population differ by age, weight, gender, and height. Tailored digital twins were developed using these correlated, individualized parameters to propose a personalized therapy for each patient. It was shown that the patients of different ages, weights, and gender have significantly different fentanyl blood uptake, plasma fentanyl concentration, pain relief, and ventilation rate, which means the same therapy will not reach the same result for the patients. Therefore, we included the virtual patients’ response to the treatment, namely pain relief, in the digital twins. We enabled these digital twins of each patient to adjust in-silico the therapy in real-time to have more efficient pain relief. By implementing digital-twin-assisted therapy, the average pain intensity decreased by 16% compared to conventional therapy. The median of time without pain increased by 23 hours over a 72-hour period. Therefore, the digital twin can be successfully used to assist in individual control of the transdermal therapy to reach higher pain relief and maintain a steady pain relief throughout the therapy.

## 1 INTRODUCTION

Fentanyl is a strong synthetic opioid that is 50 to 100 times more potent than morphine and 30 to 50 times more potent than heroin [1]–[3]. It has high lipophilicity, allowing it to quickly pass the blood-brain barrier [4]. Fentanyl transdermal patches are being extensively used for severe pain management, including cancer-induced pain [4], [5]. Fentanyl therapy has a narrow therapeutic range [6]. Therefore, keeping the fentanyl concentration in plasma in this window is crucial to reach the therapeutic effect while avoiding its toxicity. Clinical results of fentanyl transdermal therapy show that a high interindividual variability occurs in response to the therapy [7]. As such, there is a different uptake and response for each patient. Therefore, the therapeutic transdermal dose must be adapted for every patient. Due to the different physiological features of patients, their cause of pain, the intensity of the pain, and concomitant diseases, every patient needs a different dose of fentanyl. However, it is challenging to try different therapies with transdermal fentanyl therapy to find optimum therapy for each patient. Such trial and error are discomforting and dangerous for the patient and require a higher time commitment from the medical doctor.

By implementing the experimental and clinical data, and involving physics, physics-based modeling of the drug uptake can predict the patient’s plasma concentration and clinical response to different therapies. Via this approach, we are able to examine different approaches to modify it based on the patient’s needs in a shorter time without putting the patient in danger or discomfort. Numerous studies were conducted in order to develop a mathematical or computational model to predict the outcome of therapies for the patient [8]–[13]. These studies focus on different administration routes for medicinal products, such as oral, pulmonary, parenteral, and transdermal routes. These models for transdermal drug delivery are divided into two groups: 1. The studies focused on the drug’s transport from the patch through the skin at different scales [14]–[21], 2. The studies are focused on pharmacokinetics and pharmacodynamics modeling of fentanyl transport in the body [22]–[25]. However, simulations that cover the whole process of transdermal drug uptake, distribution and metabolization, clinical effect, and even patient feedback are rarer [26]. In order to reach this aim, we developed a physics-based digital twin by including virtual patient feedback[26]. The digital twin is a virtual presentation of the real-world patient that contains all his/hers organs and processes, and it is connected to the real world by sensor data or patient feedback. The drug uptake model in skin, pharmacokinetics, and pharmacodynamics model was included in this digital twin. The digital twin was used to simulate the transdermal fentanyl therapy for virtual patients of different ages. However, other patient physiology features besides age, such as weight and gender, were not considered, although they significantly affected therapy outcomes. The other patient’s characteristics must be considered in a modification of the digital twin.

In this study, a virtual population containing 3000 virtual cancer patients was generated based on sample data. The members of this population differ from each other by age, gender, weight, and height. In order to evaluate the outcome of fentanyl transdermal therapy for each patient, a physics-based digital twin was developed. The model parameters of this digital twin were modified based on patient physiology and considering inter-individual variability. The physics-based digital twin contains three main parts: (I) drug uptake model to predict the absorption of the drug from the patch through the skin to reach the blood circulation, (II) pharmacokinetics model to predict the fentanyl concentration in plasma, and (III) pharmacodynamics model to predict the two clinical effects of fentanyl, which are pain relief (therapeutic effect) and reduction in ventilation rate (adverse effect). The used parameters in these models were modified based on the patient’s physiology and inter-individual variability. The digital twin proposes a therapy for each individual to relieve the pain efficiently and avoids hypoventilation based on the calculated pain level compared to the targeted pain intensity.

## 2 MATERIALS AND METHODS

### 2.1 Digital twin

#### 2.1.1 Drug uptake model

The drug uptake model simulates the drug diffusion in the transdermal patch and skin layers (stratum corneum, viable epidermis, and upper part of dermis). The thickness of the skin layer was modified for each patient, as this was dependent on patient physiology, such as gender, weight, and age. The geometric dimensions for the patch were based on Duragesic® fentanyl patches. The skin and patch have a square shape in the model, namely as wide as the transdermal patch. We assumed that there was perfect contact between the patch and the skin. The implemented geometry in this study is shown in Figure 1.

**Figure 1.**
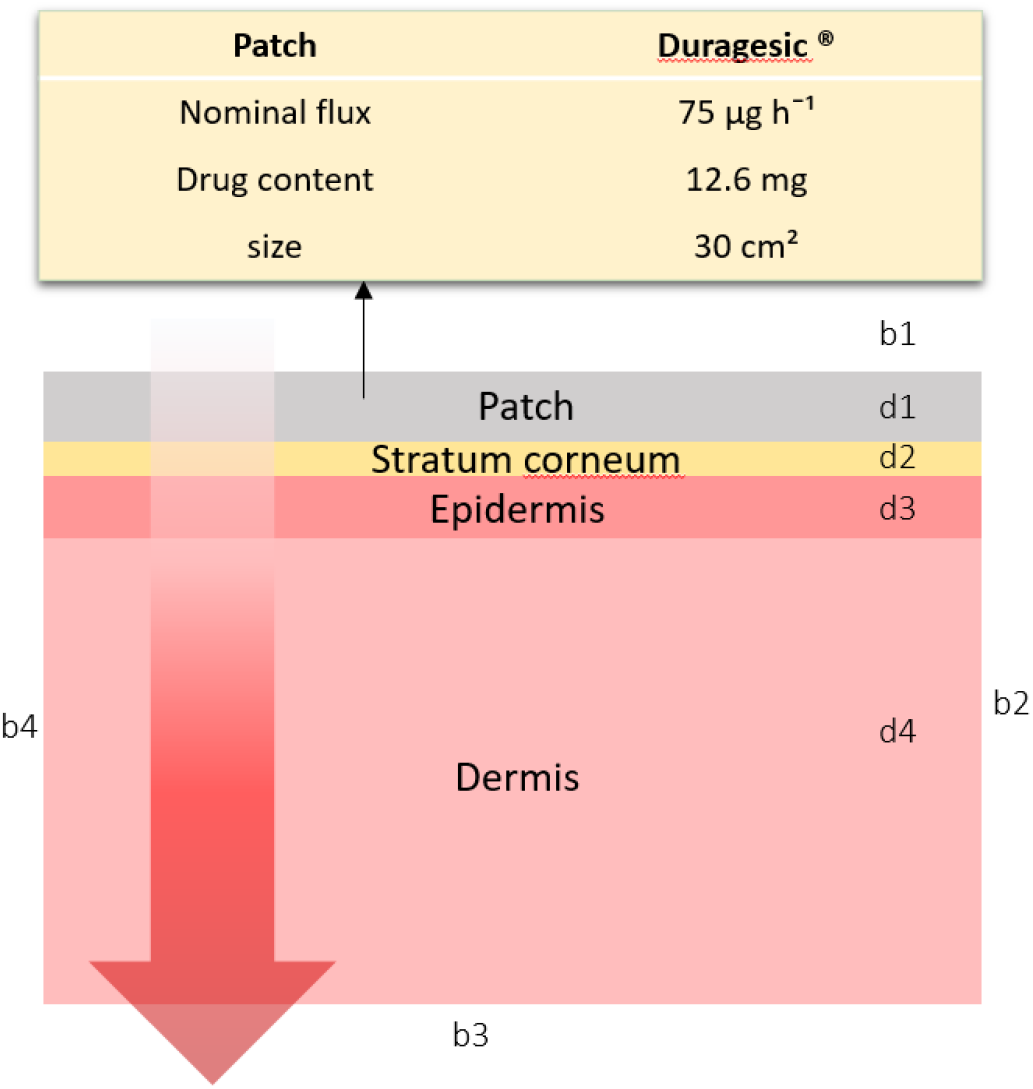
Implemented geometry of skin and patch for drug uptake model; Boundaries b1, b2, and b4 have no flux; however, fentanyl is absorbed by the blood circulation system in the b3 boundary. In this model, the patch’s thickness is 50.8 µm, the stratum corneum’s thickness varies in the range of 12-27 µm, the thickness of the viable epidermis is between 15 to 51 µm, and the thickness of the upper part dermis is between 153 to 374 µm.

##### 2.1.1.1 Computational system configuration

##### 2.1.1.2 The governing equation

The drug penetration from the patch through the skin was modeled by transient diffusion which is described by Fick’s second law (Equation 1).

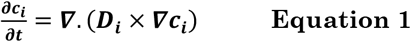

where c_i_ and D_i_ are concentration [ng/ml] and diffusion coefficient [m^2^/s] of fentanyl in domain i. At the interface of each two layers, partitioning should be considered. The partition coefficient at each interface is mentioned in Equation 2.

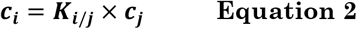

where K_i/j_ is the partition coefficient between the two domains i and j. The partition coefficient is the concentration ratio of one solute at the interface of two immiscible phases. Due to the partition coefficient, the concentration of fentanyl at the two sides of the interface of the layers might be different. This discontinuity is inconvenient to solve computationally. To solve this problem, the drug’s potential Ψ_i_, as described in Equation 3, it was used instead of the concentration, as it is continuous throughout the layers.

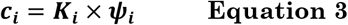

where K_i_ is drug capacity in domain i, and it is related to partition coefficient as: K_i/j_ = K_i_/K_j_ (as a result of continuity of Ψ_i_ over the whole domain).

Fentanyl penetrates through the epidermis and reaches the dermis. In the dermis layer, the drug will be taken up by capillary vessels at different lengths of the dermis. Besides diffusion, other physical mechanisms such as advection will play a role. In this study, we only modeled the diffusion process. Additionally, the capillaries will absorb the fentanyl in the upper part of the dermis, and all of the drug or part of it will be absorbed before reaching its bottom. To obtain the same drug uptake through the skin as the clinical studies, we used an equivalent dermis thickness, which is smaller than the real dermis thickness. The details of this assumption and modification and validation of drug uptake model are mentioned in our previous studies [26]–[28].

##### 2.1.1.3 Boundary and initial conditions

At the top boundaries (Figure 1, b1) and for the peripheral boundaries (Figure 1, b2& b4), we assumed no flux of fentanyl. The drug in the patch contains 12.6 mg of fentanyl (Figure 1, d1). At the bottom boundary (Figure 1, b3), the concentration of fentanyl is taken equal to the concentration of fentanyl in blood. This concentration is obtained from the pharmacokinetic model. The initial concentration of fentanyl in the skin layers (Figure 1, d2, d3& d4) is equal to zero, as there is no drug in the skin when applying the patch. This assumption implies that the patch is always applied at a new location.

##### 2.1.1.4 Pharmacokinetics modeling

In this study, a physiologically-based pharmacokinetic model was used to predict the concentration of fentanyl in the blood plasma. Due to the complexity of the human body, different organs were lumped together into PK compartments [26]. We considered five compartments: (I) the central compartment, which contains blood and lungs (Equation 4); (II) the rapid equilibrated compartment, which includes the brain, heart, skin, and kidneys (Equation 5); (III) the slow equilibrated compartment, which contains muscle, fat tissue, and the carcass (Equation 6); (IV) gastrointestinal compartment which includes gut, spleen, and pancreas (Equation 7); (V) hepatic compartment where the metabolization happens and which only includes the liver (Equation 8). The concentration of fentanyl in these compartments is evaluated by following ordinary differential equations:

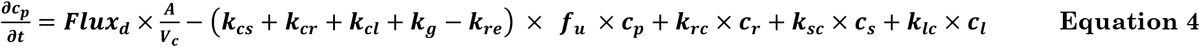

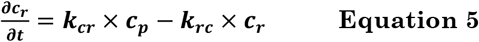

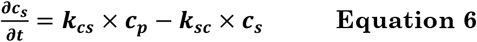

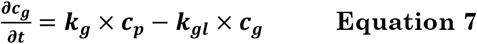

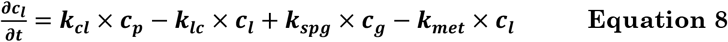

The parameters of these equations are mentioned in Table 2. In this set of equations, c_p_, c_r_, c_s_, c_g_, and c_l_ are the concentration of fentanyl in the central, rapid equilibrated, slow equilibrated, gastrointestinal and hepatic compartments, respectively. f_u_ is the unbound fraction of fentanyl in the central compartment. k_ij_, k_met_, and k_re_ are the first-order equilibrium rate constants for inter-compartmental clearance, metabolization, and renal clearance, respectively. In our previous study, the pharmacokinetic model was validated[26].

#### 2.1.2 Pharmacodynamics model

There is a delay between the concentration of fentanyl in the blood plasma and the corresponding clinical effects on pain relief and breathing rate [29]. To account for this delay, a virtual compartment is defined, called the effect compartment. The concentration of fentanyl in the effect compartment related to the central compartment was calculated by Equation 9.

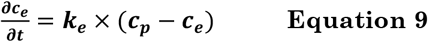

As fentanyl is a small lipid-soluble drug, it will rapidly penetrate through the blood-brain barrier from the blood circulation system and bind to opioid receptors in the central nervous system [30]. Through this mechanism, fentanyl produces pain relief. We used the visual analog scale (VAS), which is a pain scale between 0 (indicates no pain) and 10 (indicates the worst possible pain), to evaluate the pain relief. Additionally, fentanyl will bind to the opioid receptor on chemoreceptors in the brain stem, which are responsible for detecting the carbon dioxide level in blood and stimulating breathing. This binding will reduce the sensitivity of carbon dioxide levels and may lead to stopping breathing in extreme cases. A sigmoidal model models the reduction of the ventilation rate and VAS pain score as a function of fentanyl concentration in the effect compartment (Equation 10).

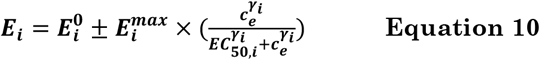

Where, 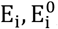, and 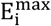 are the intensity of the pharmacologic, baseline, and maximum reachable effect, respectively. EC_50,i_ is the concentration related to the response halfway between the baseline and maximum possible effect and γ_i_ is the Hill coefficient. In our previous study, the parameters for the pharmacodynamics model for pain relief were derived based on experimental data[26].

### 2.2 Generation of the virtual population

#### 2.2.1 Sample data of cancer patients

Ten women and ten men with age 40 to 68 with different tumor sites were studied by Zech et al. [31]. All the involved patients used fentanyl transdermal therapy for chronic pain management. The patient’s physiological features are mentioned in Figure 2, including age, gender, weight, and height. These parameters are cross-correlated. To explore the correlation between these physiology features, the correlation test was implemented on this set of data, and its result is shown in Figure 2. This result shows a strong correlation (correlation coefficient (C.C.)=0.87) between gender and height. However, it should be clarified that the value for gender is considered Boolean in this set of data. The value for the male gender is one and for the female gender is zero. Therefore the positive correlation coefficient of 0.87 represents the higher height between male genders as is expected. The correlation test result also represents a weak correlation between gender and weight (C.C.=0.43) and weight and height (C.C.= 0.38).

**Figure 2.**
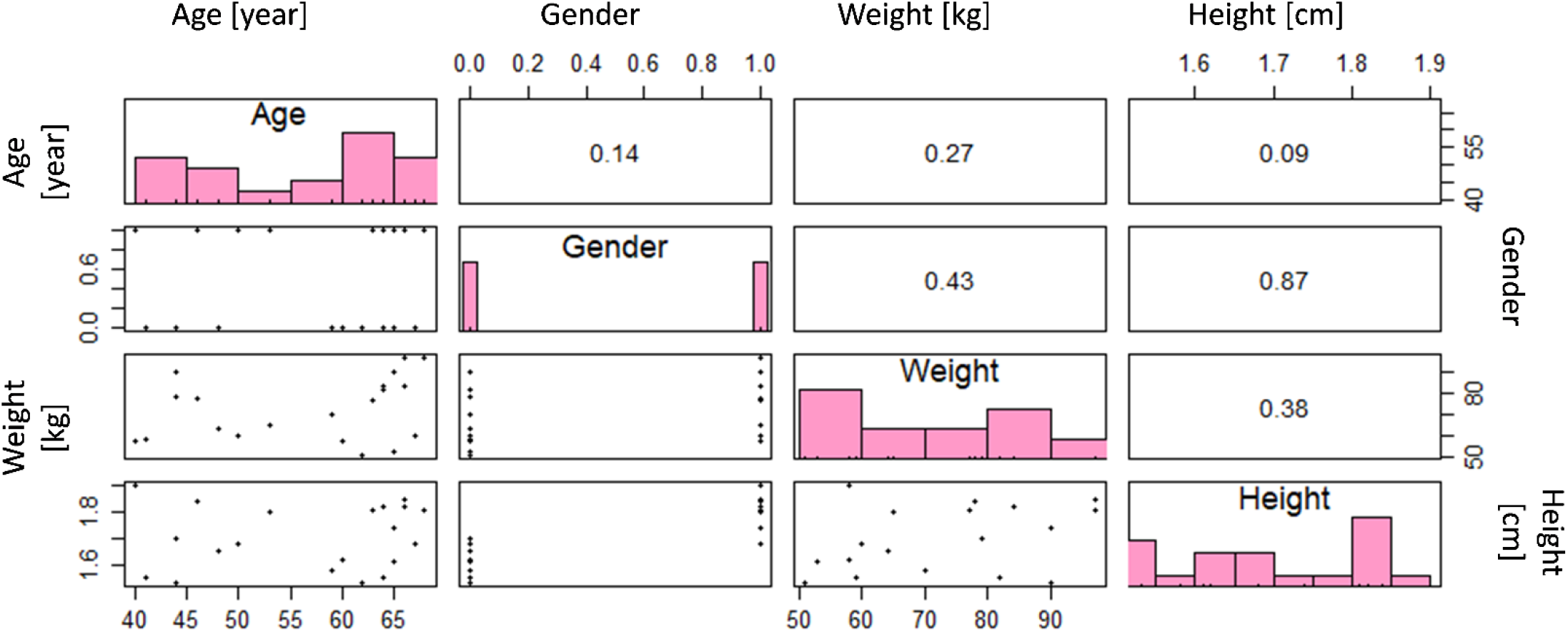
Pearson correlation coefficient between each two sample data parameters and the distribution of the values for these parameters.

#### 2.2.2 Virtual population

In this study, we developed a virtual population of patients from sample data to examine the performance of digital twins for patients with a wider range of physiological features. Members of this population differ from each other based on their age, gender, weight, and height. In reality, these physiological features of patients are correlated, as shown in Figure 2. Therefore, our virtual patients should have correlated features in the same way. We used the Markov chain Monte Carlo (MCMC) method to produce the virtual population with realistic feature combinations for each patient. MCMC predicts the posterior distribution by sampling from the joint distribution of likelihood and prior distribution. This method uses the sample demographic data of cancer patients (N=20) as a likelihood function. The prior distribution for age, weight, and height is considered normal, and gender is considered a uniform distribution. By using the Gibbs sampling method, the posterior distribution (N_burn_=20000 and N_keep_=3000) was evaluated. The demographic characterization of virtual patients of the generated population (N=3000) was obtained from this joint distribution. In Figure 3, the generated virtual population is shown. By comparing the correlation coefficients between predicted features for virtual population and sample data, we find that the MCMC method was able to mimic the correlation between sample data. However, the distribution of parameters in the sample data and population data are visually different due to the limited number of cases in the sample data. The virtual population members are chosen based on sample data to reach a population with physiological features compatible with sample data of real patients. The summary of sample data and population data are shown in Table 1.

**Table 1.** Patient characteristics for sample data and population

|  | Sample data [31] | Population data |
| --- | --- | --- |
| Number | 20 | 3000 |
| Age [year] | 56.75 (SD=9.82) | 56.70 (SD=4.9) |
| Weight [Kg] | 72.8 (SD=14.8) | 72.82 (SD=7.57) |
| Gender (M/F) | 10/10 (50% male and 50% female) | 1512/1488 (50.4% male and 49.6% female) |
| Height [m] | 1.70 (SD=0.12) | 1.70 (SD=0.06) |

**Figure 3.**
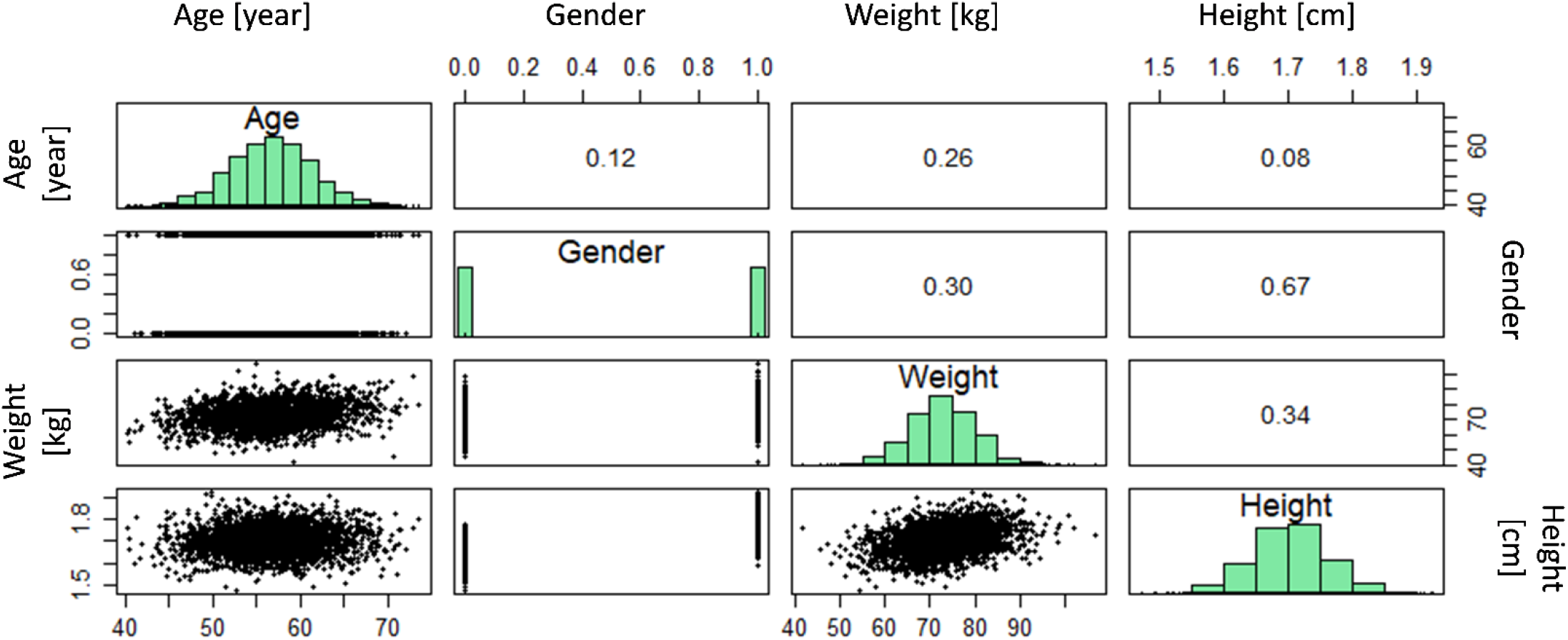
Pearson correlation coefficient between each two population data parameters and the distribution of the values for these parameters.

#### 2.2.3 Estimation of model parameters

The patient’s physiology changes the skin properties, volume of the organs, blood flow, glomerular filtration rate, metabolization, and opioid tolerance of the patient. By using the patient’s individual physiological features, we can predict the input parameters for the model. It should be noted that despite these equations being obtained from experimental data, as a result of inter-individual variability, different values for these parameters correspond to patients with the same age, gender, weight, and height. To account for this inter-individual variability, the calculated value was modified by the exponential of a normal random number with mean=0 and SD= 0.1. By including this random factor, 95% of the evaluated parameters are in the range of ±20% of the evaluated values. The overall dependency of the model parameter on patient physiology is mentioned in Table 2. As mentioned earlier, if the gender of the patient is male, the value for the gender parameter is equal to 1 and for the female is 0.

**Table 2.**
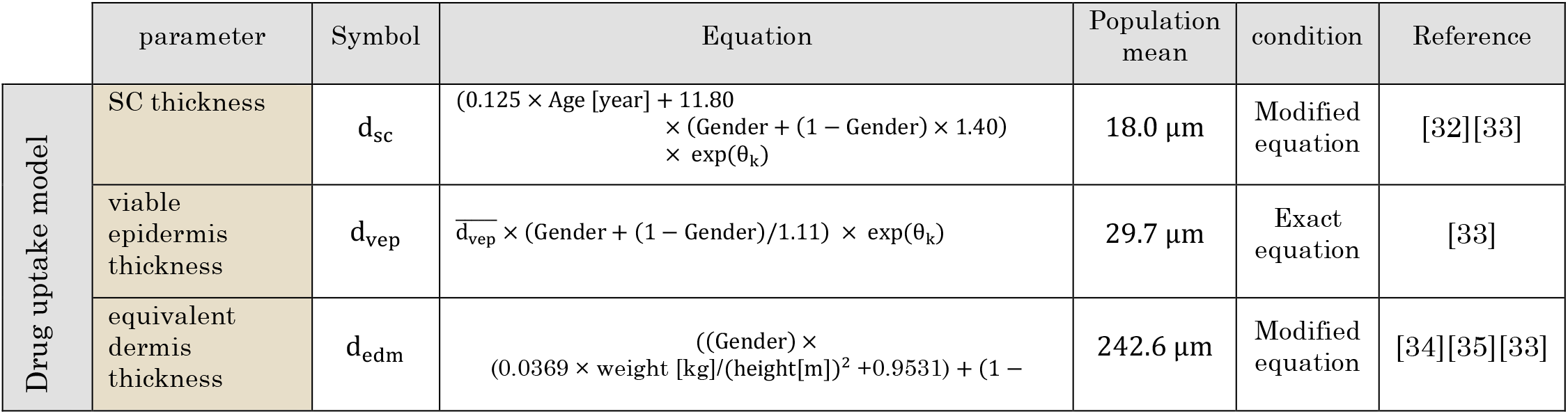
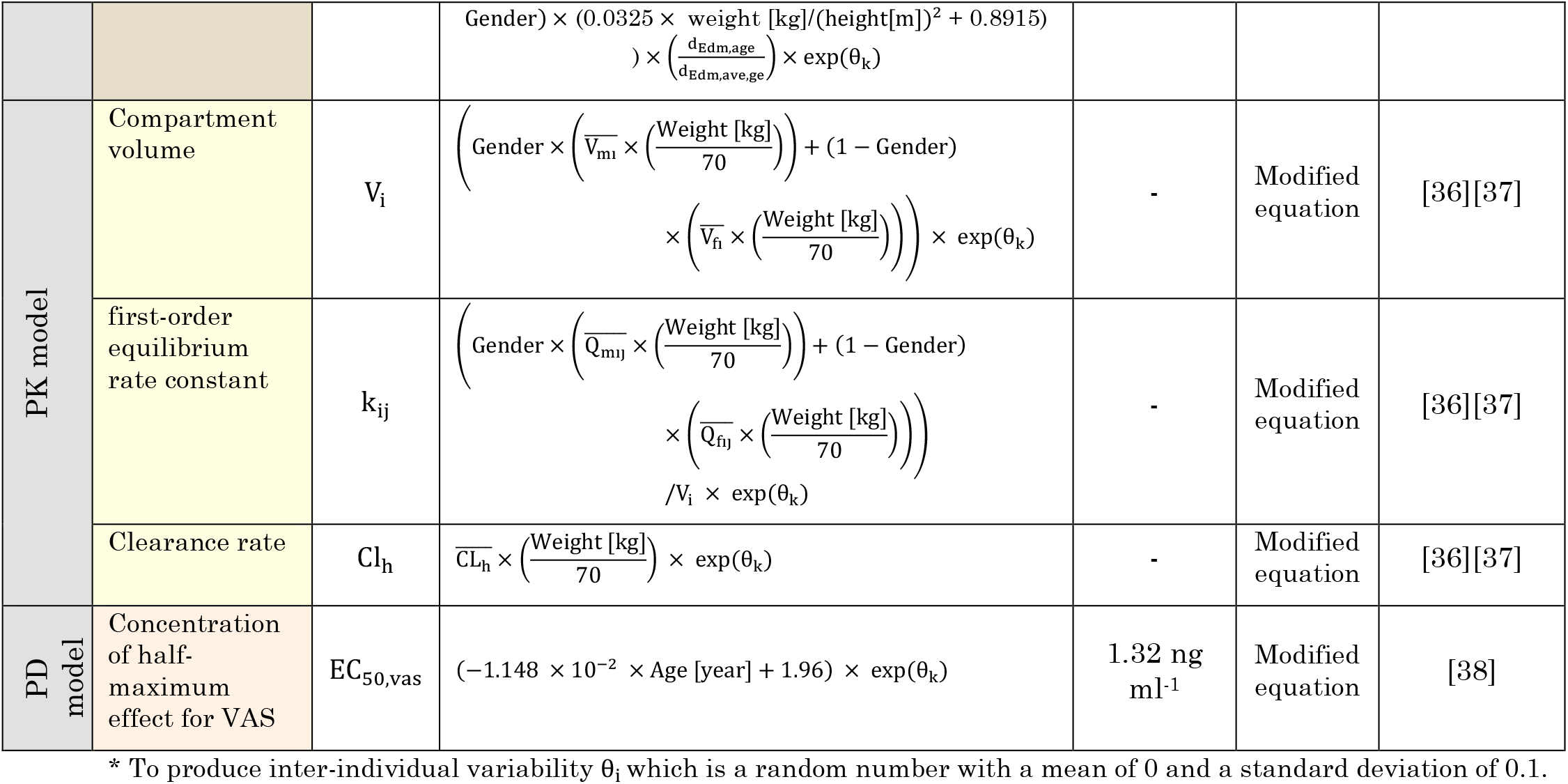
The dependent model parameters to patient physiological features

Based on Table *2*, skin layer thickness, compartment volumes, rate constants, clearance rate, and the concentration of half maximum effects for pain relief were directly calculated based on physiological features. The rest of the model parameters in this study were considered independent of the patient’s physiological features and are in Table 3. In real life, these parameters might depend on the patient’s physiology. However, for the four features considered in this study, based on our best knowledge, there is no proposed experimental model to calculate these parameters based on these four physiological features. By considering both parameters, dependent and independent of patient physiology, all input parameters change between the patients.

**Table 3.**
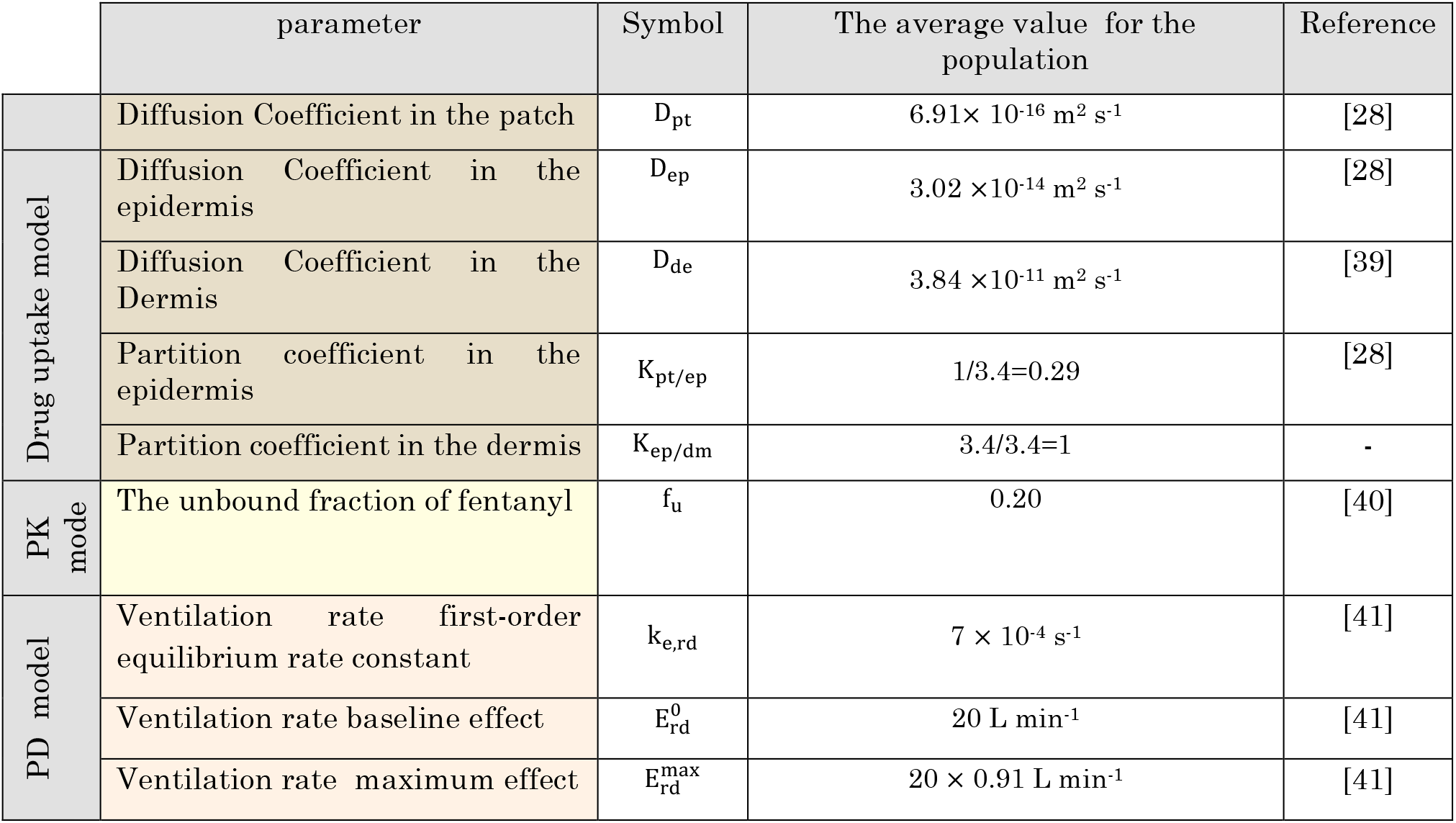
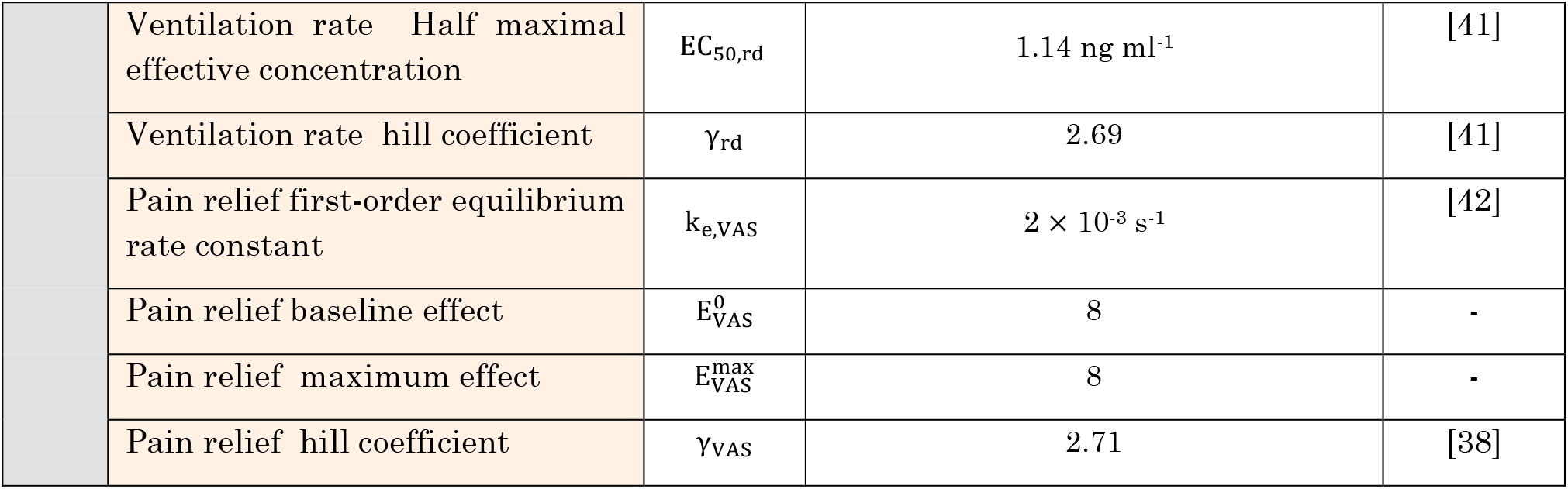
the independent model parameters to patient physiological features

|  | parameter | Symbol | The average value for the population | Reference |
| --- | --- | --- | --- | --- |
| | Diffusion Coefficient in the patch | $D_{pt}$ | $6.91 \times 10^{-16} \text{ m}^2 \text{ s}^{-1}$ | [28] |
| Drug uptake model | Diffusion Coefficient in the epidermis | $D_{ep}$ | $3.02 \times 10^{-14} \text{ m}^2 \text{ s}^{-1}$ | [28] |
| | Diffusion Coefficient in the Dermis | $D_{de}$ | $3.84 \times 10^{-11} \text{ m}^2 \text{ s}^{-1}$ | [39] |
| | Partition coefficient in the epidermis | $K_{pt/ep}$ | 1/3.4=0.29 | [28] |
| | Partition coefficient in the dermis | $K_{ep/dm}$ | 3.4/3.4=1 | - |
| PK model | The unbound fraction of fentanyl | $f_u$ | 0.20 | [40] |
| PD model | Ventilation rate first-order equilibrium rate constant | $k_{e,rd}$ | $7 \times 10^{-4} \text{ s}^{-1}$ | [41] |
| | Ventilation rate baseline effect | $E_{rd}^0$ | 20 L min<sup>-1</sup> | [41] |
| | Ventilation rate maximum effect | $E_{rd}^{\text{max}}$ | $20 \times 0.91 \text{ L min}^{-1}$ | [41] |

|  |  |  |  |
| --- | --- | --- | --- |
| Ventilation rate Half maximal effective concentration | $EC_{50,rd}$ | 1.14 ng ml<sup>-1</sup> | [41] |
| Ventilation rate hill coefficient | $\gamma_{rd}$ | 2.69 | [41] |
| Pain relief first-order equilibrium rate constant | $k_{e,VAS}$ | $2 \times 10^{-3} \text{ s}^{-1}$ | [42] |
| Pain relief baseline effect | $E_{VAS}^0$ | 8 | - |
| Pain relief maximum effect | $E_{VAS}^{\max}$ | 8 | - |
| Pain relief hill coefficient | $\gamma_{VAS}$ | 2.71 | [38] |

### 2.3 Spatial and temporal discretization

The grid sensitivity analysis by using Richardson extrapolation was done over the skin layers and fentanyl patch based on the flux out of the dermis, in which spatial discretization error was considered 0.1%. As a result of this grid sensitivity analysis, about 900 quadrilateral grids were built. However, this number varies from case to case as the thickness of the skin’s layer differs between cases. In some cases, almost 100 quadrilateral grids were sufficient. In order to increase the numerical accuracy, the accumulation of grids near the interfaces is more. To define the time steps, sensitivity analysis based on maximum fentanyl concentration was done, which defined a maximum time step of 6 hours; however, to reach a higher temporal resolution, time steps were assigned as 1 hour.

### 2.4 Numerical implementation and simulation

COMSOL Multiphysics version 5.4 was used in this study to solve the diffusion process of fentanyl from the patch through the skin in the mechanistic model, the distribution of fentanyl in the human body in the pharmacokinetics model, drug’s effect in the pharmacodynamics model. We implemented MUMPS (MUltifrontal Massively Parallel sparse direct Solver) solver scheme in our simulation. A partial differential equation (PDE) interface solved the diffusion process of fentanyl in the mechanistic model was solved by a partial differential equation (PDE) interface. To take to account the distribution, elimination, and metabolization of fentanyl, the boundary ordinary differential equation (ODE) was used. In the pharmacodynamics model, the concentration of fentanyl in the effect compartment was calculated by the PDE interface, and the boundary probe calculated the drug’s effect. To apply the change of the patch and skin location during the therapy, the event interface was used. The population generation was done in RStudio by using the “mixAK” package. Analyzing the sample data, calculating the posterior distribution, generating the virtual patients’ characteristics, calculating the model parameters, and analyzing the result of digital twins are done in RStudio.

### 2.5 Proposed therapy by the digital twin

The tailored physics-based digital twin predicts the outcome of fentanyl transdermal therapy for the patient at any moment of the treatment. Based on the summary of product characteristics (SmPC), which we will call “conventional therapy” in this study, the fentanyl transdermal patch needs to be changed every 72 hours. After applying the patch, fentanyl accumulates in a depot in the skin, and it is gradually released into the blood circulation, and after reaching its peak in blood, the concentration in the blood decreases as the amount of drug in the patch decreases. As a result of the lower concentration, the analgesic effect decreases too. Patients who don’t reach sufficient analgesic effect within 72 hours intervals can switch to 48 hours intervals [43]. This prescription is similar for every patient as there is a limited number of alternative approaches[26]. However, by using the predicting capability of the digital twin, we can propose a dynamic application time for each patch based on patient needs. In this regard, every 8 hours, the digital twin considers the condition for changing the patch. If the pain intensity is above target (in this case, VAS pain score equal to 3), the patient needs to change the patch. The reason for putting a limit of 8 hours is to avoid changing the patch too frequently, which may lead to an intensive increase in concentration in plasma and drug waste. Additionally, there is a 1 to 2 hours of a time lag between the application of fentanyl transdermal patch and reaching the blood circulation [44]. Based on our previous study, the time to reach half maximum concentration is about 8 hours [26]. In this way, the digital twin gives enough time to evaluate the patient’s pain relief. The general approach of digital-twin-assisted therapy is shown in Figure 4.

**Figure 4.**
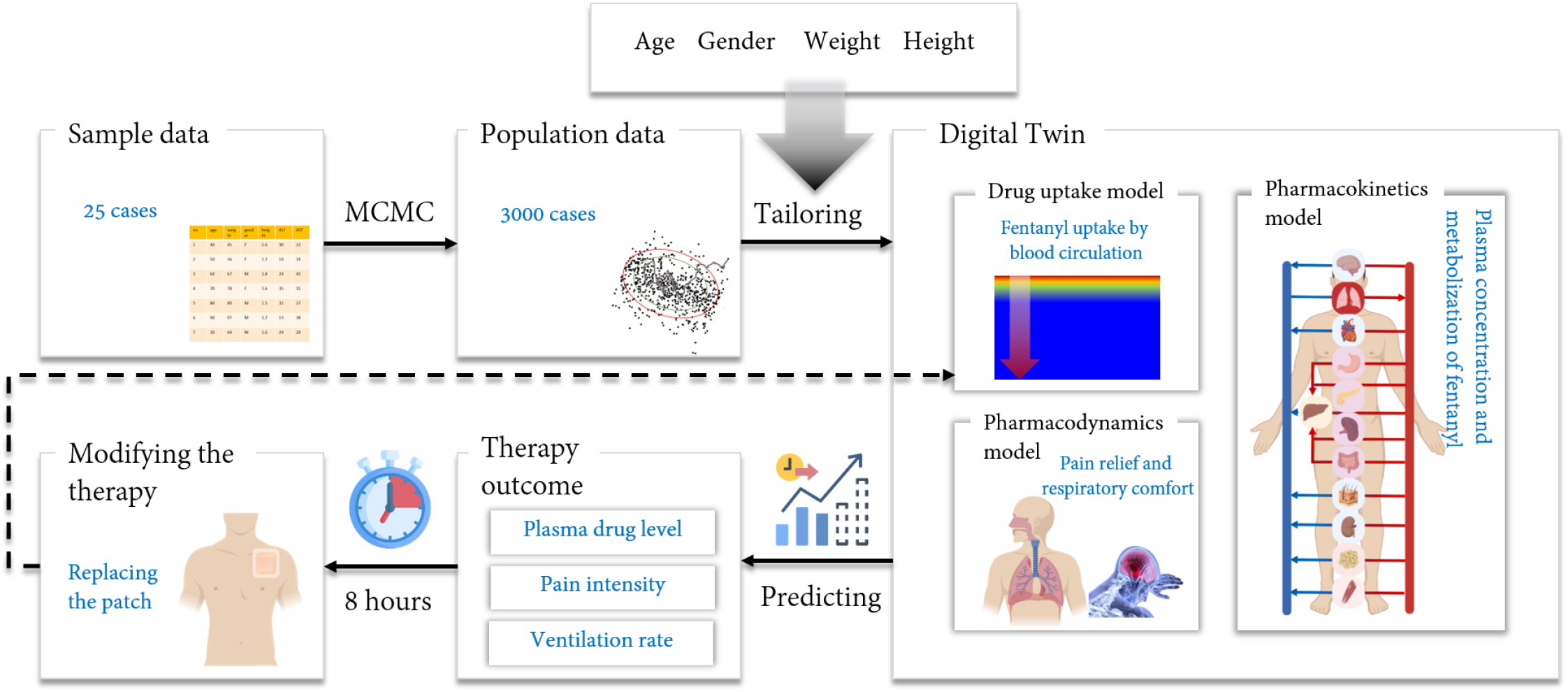
The overall structure of tailored digital twin for each member of the virtual population

## 3 RESULTS and DISCUSSION

### 3.1 Conventional therapy

The conventional therapy was applied to all virtual patients over three days. Three thousand virtual patients implemented a Duragesic ® fentanyl patch with a nominal flux of 75 µg h-1 over 72 hours based on the SmPC. The outcome of therapy was evaluated based on three output parameters: fentanyl concentration in plasma, pain intensity, and ventilation rate. This result is shown in Figure 5. In Figure 5/a-c, the outcome based on patient age is represented, which shows no distinct trend in the effect of age on fentanyl concentration in plasma, pain intensity, or ventilation. This implies the interindividual variability, and the effect of characteristic features of the patient overshadows the effect of age. The previous study showed that the age of the patient drastically affects the therapy’s outcome[26]. By comparing these two studies, we conclude that pain relief will increase for an individual by aging with the same amount of fentanyl. Due to the high impact of other characteristics of the patient, with solely the patient’s age, we cannot predict the therapy outcome. Figure 5/d-f shows the outcome of therapy versus weight. Based on the result, by increasing weight, the concentration in plasma drops, which leads to lower pain relief and simultaneously lower side effects, which is the reduction in ventilation rate. The reduction of fentanyl concentration in plasma by increasing weight is due to the larger volume of each compartment; the drug will be diluted. Therefore, weight has a key impact on pain relief, which is the main aim of the therapy. In the next step, we studied the effect of height on the outcome of the therapy. This result is demonstrated in Figure 5/g-I, which shows that the concentration in plasma reduces by increasing height. As a result of the reduction in plasma concentration, pain relief reduces as well. However, based on Figure *2* Figure *3*, higher height is correlated to higher weight, and the reduction in plasma concentration is due to higher weight, not necessarily higher height. Here in these three graphs, we see a gap between genders due to the height gap between the genders. Nevertheless, standard fentanyl therapy clearly is not gender-neutral.

**Figure 5.**
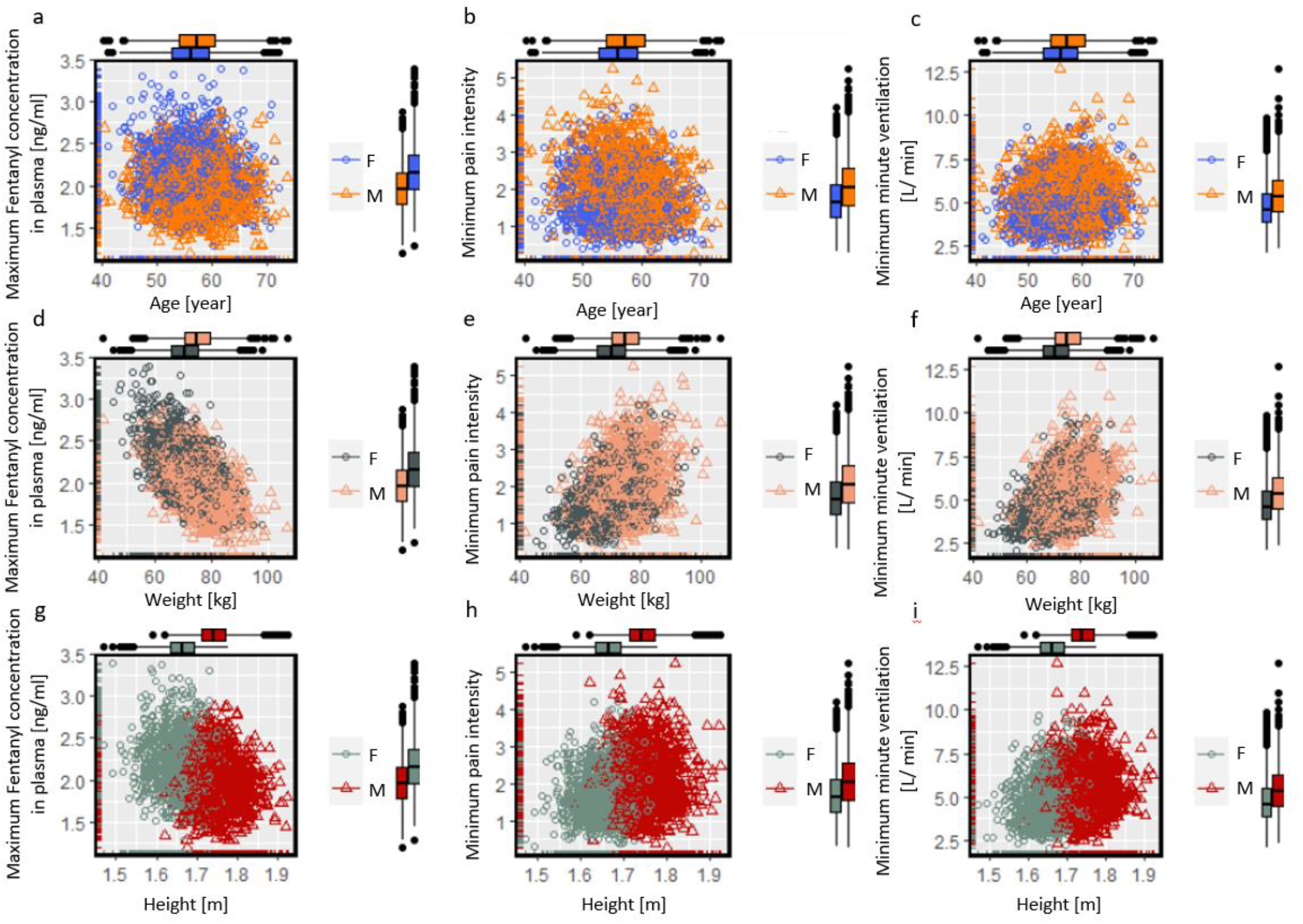
Result of conventional fentanyl transdermal therapy over 72 hours for: a. maximum fentanyl concentration in plasma based on age, b. minimum pain intensity based on age, c. minimum ventilation rate based on age, d. maximum fentanyl concentration in plasma based on weight, e. minimum pain intensity based on weight, f. minimum minute ventilation based on weight, g. minimum fentanyl concentration in plasma based on height, h. minimum pain intensity based on height, i., minimum minute ventilation based on height.

### 3.2 Digital-twin-assisted therapy

As mentioned in section 2.5, digital twins predict the pain intensity and how the treatment influences it. Based on the intensity of the pain, the digital twin can propose changing the patch if the pain intensity is higher than the target (VAS pain score 3). The digital twin is able to calculate the pain intensity at any moment and update the therapy based on that. However, based on our previous study, it takes about 8 hours for a fentanyl patch to reach half of the maximum contraction in plasma. Therefore, we put a limit of 8 hours for the checking points to update the therapy. At first, for the patient number one (age=58 years, Male, Weight=74.2 [kg], and Height=1.73[m]), we explored the effect different checking points at every 2, 4, 6, 8, 12, 24, 36, and 72 hours. We did not consider frequencies shorter than 2 hours, as the time lag of fentanyl in reaching the blood circulation is about 2 hours [44]. In Figure 6/a, the profile of pain relief for different checking points is shown and based on Figure 6/b, except for the checking point every 24 and 72 hours, and the average pain intensity is lower than 3. The checking point at every 36 hours has better pain relief than 24 hours because this specific patient has higher pain intensity around 36 hours after therapy. Additionally, based on Figure 6/c, for all the checking points duration every 24 and 72 hours, the time without pain was more than 48 hours (more than 2/3 of the therapy duration). As shown in Figure 7/d, except for the checking point every 72 hours, all other cases pass the plasma concentration threshold; however, the criteria for breathing rate are met by all the cases (shown in Figure 6/e). In Figure 6/f, the number of the needed patch by the first patient, by considering different checking points. Considering the pain relief performance while avoiding excessive patches, the four checkpoints of 6, 8, 12, and 36 hours match our target in pain relief and minute ventilation. Other virtual patients might need checking more or less frequently based on their therapy outcome. However, we chose 8 hours interval as it matches the time to reach half of the maximum plasma concentration in our previous study, and it is frequent enough that the patient does not remain in pain.

**Figure 6.**
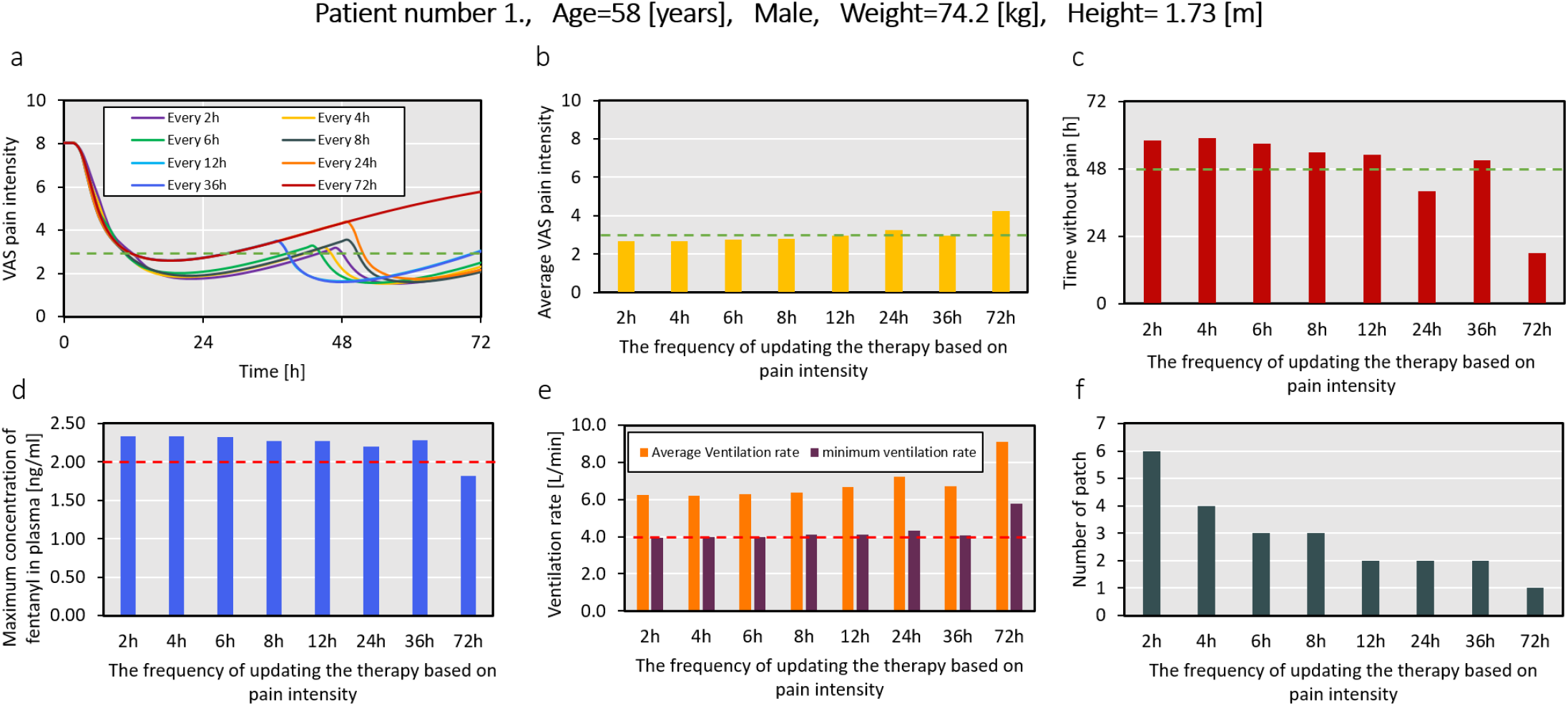
a. VAS pain intensity for patient number one, during 72 hours, by updating the treatment at different frequencies; b. the average pain intensity for the patient as a result of varying updating points and c. the subsequent time without pain (VAS pain intensity below 3); d. the maximum concentration of fentanyl in plasma, and e. the average and minimum ventilation rate throughout the therapy by considering the different frequencies of updating the therapy; f. the number of needed patch for each scenario.

**Figure 7.**
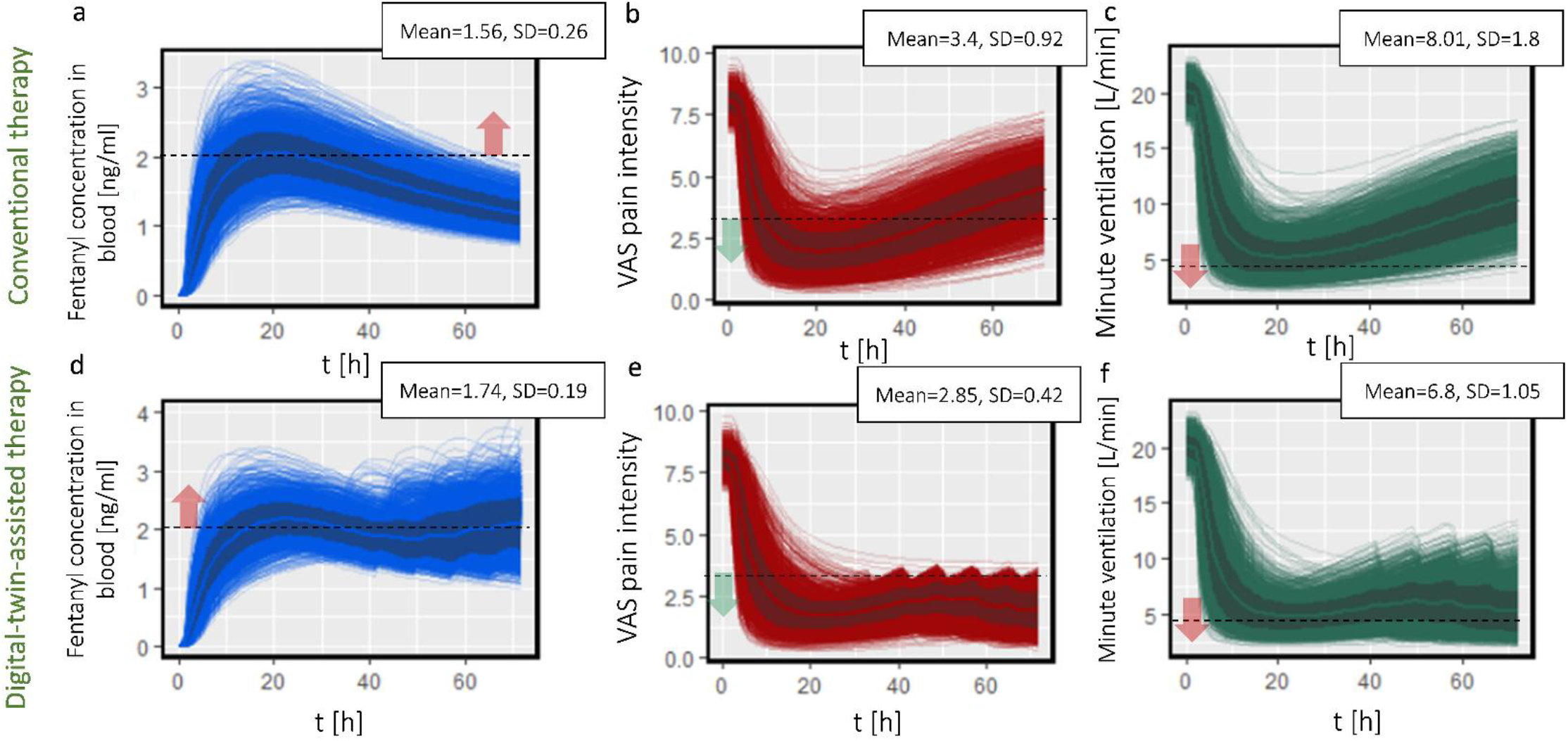
a. fentanyl concentration in plasma profile during conventional therapy, b. VAS pain intensity during conventional therapy, c, minute ventilation rate during conventional therapy, d. fentanyl concentration in plasma profile during digital-twin-assisted therapy, e. VAS pain intensity during digital-twin-assisted therapy, f. minute ventilation rate during digital-twin-assisted therapy in 72 hours for all virtual patients.

The concentration of fentanyl in plasma for all the virtual patients during conventional therapy versus time is shown in Figure 7/a. As shown in the graph for a large number of patients, the fentanyl concentration in plasma reaches the upper threshold during the therapy. It should be noted that the threshold of 2 ng ml^-1^ [45] is an average for all patients. This number in other studies is 3 ng ml^-1^ [46]. As the concentration of fentanyl in plasma after reaching the maximum reduces, the patient’s pain intensity starts to increase as well, as shown in Figure 7/b. More than 15% of the patients experience minute ventilation below 4 L/min during the treatment, as shown in Figure 7/c. In the next step, we implemented digital-twin-assisted therapy for each patient. The outcome of digital-twin-assisted therapy is shown in Figure 7/d-f. By comparing the fentanyl concentration in plasma for digital-twin-assisted therapy (as it is shown in Figure 7/d) with conventional therapy, we realized the average concentration in plasma increased by 11.5% while the standard deviation decreased. This increase in concentration was necessary to keep the pain intensity below target. However, via this method, the fluctuation in concentration reduces. The result in Figure 7/e shows that the digital twin successfully kept the pain intensity for 98.8% of the patients below 3 for more than half of the duration of the treatment. However, for conventional therapy, only 57.1% of the patients have pain intensity below 3 VAS for more than half of the treatment duration. By implementing digital-twin-assisted therapy, the pain intensity decreased by 16% compared to conventional therapy, and the standard deviation decreased. A lower standard deviation corresponds to a lower deviation for pain intensity between patients. The higher fentanyl concentration in plasma reduced the average minute ventilation by 15% compared to conventional therapy.

Besides reducing the overall pain intensity, the aim of digital-twin-assisted therapy is to keep the pain below the target. In this study, we defined a parameter for the duration that the patient experiences pain lower than VAS pain score 3, called the time without pain. By implementing digital-twin-assisted therapy, the median time without pain for the population increases by 23 hours compared to conventional therapy, which is shown in Figure 8/a. The interquartile range (IQR) of time without pain for conventional therapy is 23 hours, while it is 7 hours (70% less) for digital-twin-assisted therapy. This shows that the variation of therapy’s outcome for digital-twin-assisted therapy is less than conventional therapy. In the next step, we studied the effect of patient physiology in time without pain. In Figure 8/b&f, the effect of gender in the duration that the patient has lower pain intensity is shown. During conventional therapy, the difference between the median time without pain for the male patients and the female patients is 5 hours, while this number is 40% less (3 hours) during digital-twin-assisted therapy. This shows by proposed therapy by digital twin, the effect of gender on the outcome of therapy will reduce. The time without pain corresponds to the patient’s age, weight, and height both for conventional and digital-twin-assisted therapy, as shown in Figure 8/ c-e,g-i. The correlation coefficient between time without pain and patient age through both therapies is 0.1. However, the correlation coefficient of time without pain through conventional therapy and digital-twin-assisted therapy and weight is -0.41 and -0.31, respectively. This implies that age does not affect the outcome of therapy, similar to conventional therapy. However, during digital-twin-assisted therapy, the outcome of therapy is less dependent on the patient’s weight compared to conventional therapy. Which is evaluated for the studied range of weight for virtual patients (42-107 kg), and lower or higher weights might change the results. The correlation coefficient between time without pain and patient heights through conventional therapy and digital-twin-assisted therapy is -0.15 and -0.23. However, as mentioned earlier, the patient’s height affects the model parameters via its effect on the BMI. Therefore, calculating the correlation coefficient for BMI will be -0.30 and -0.14 for conventional therapy and digital-twin-assisted therapy, respectively. By considering the result in Figure 7 and Figure 8, we conclude that digital-twin-assisted therapy successfully increased the pain relief for the patient and decreased the dependency of therapy outcome on patient physiology. With Digital-twin-assisted therapy, we are able to provide the same efficiency of pain relief for men and women, old and young patients, and patients with higher or lower BMI.

**Figure 8.**
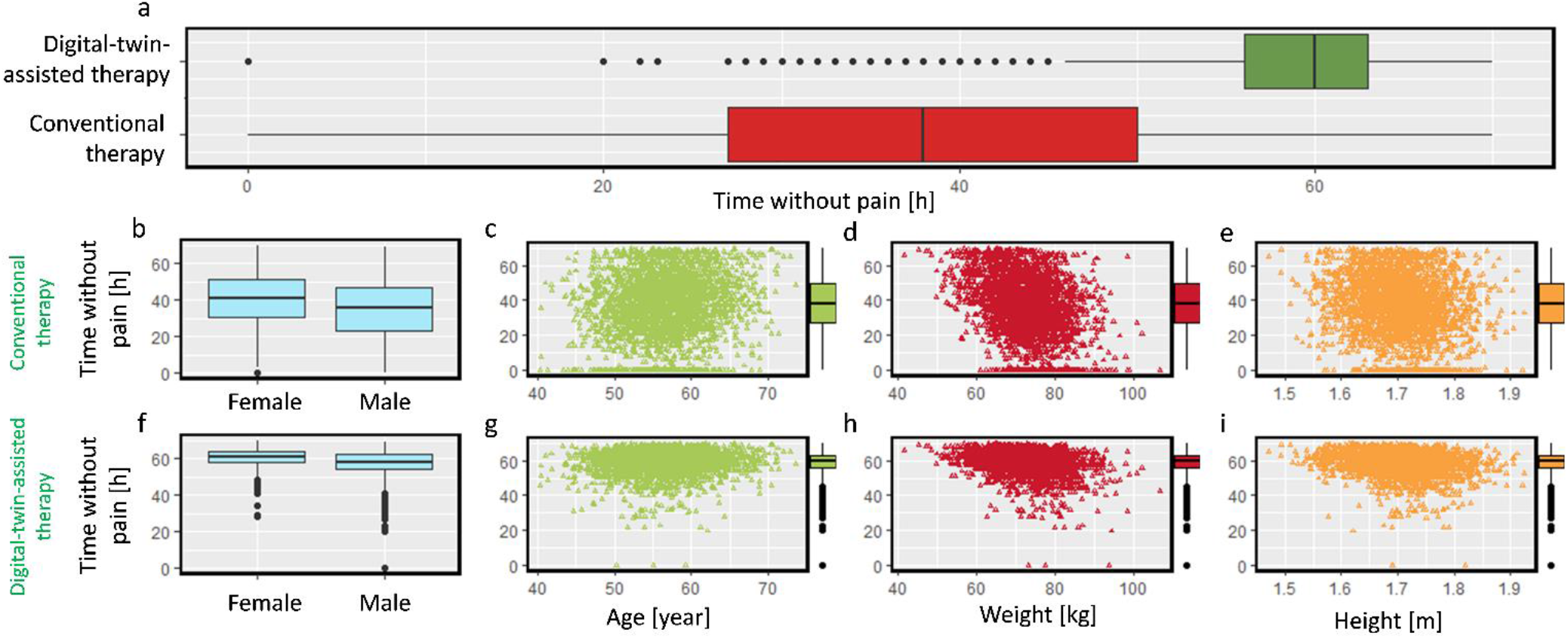
a. time without pain during 72 hours during conventional therapy and digital-twin-assisted therapy, time without pain during conventional therapy: b. based on gender, c. based on age, d. based on weight, e, based on height, time without pain during digital-twin-assisted therapy: f. based on gender, g. based on age, h. based on weight, i. based on height.

Through digital-twin-assisted therapy, each patient will use an individualized number of patches for 72 hours based on their pain intensity experience. Figure 9/ a presents the number of used patches during 72 hours based on patient weight. Based on this result, patients with higher weight, on average, will need to change the patch more frequently compared to lower weight to achieve constant pain relief. As also mentioned earlier, age does not influence the number of needed patches. By analyzing data, we realize that the majority of the patients who need to change the patch more frequently are male. This is due to the fact that, on average, male patients are taller; therefore, they tend to have a higher weight. Therefore, they proportionally need more fentanyl to keep the pain intensity below the target.

**Figure 9.**
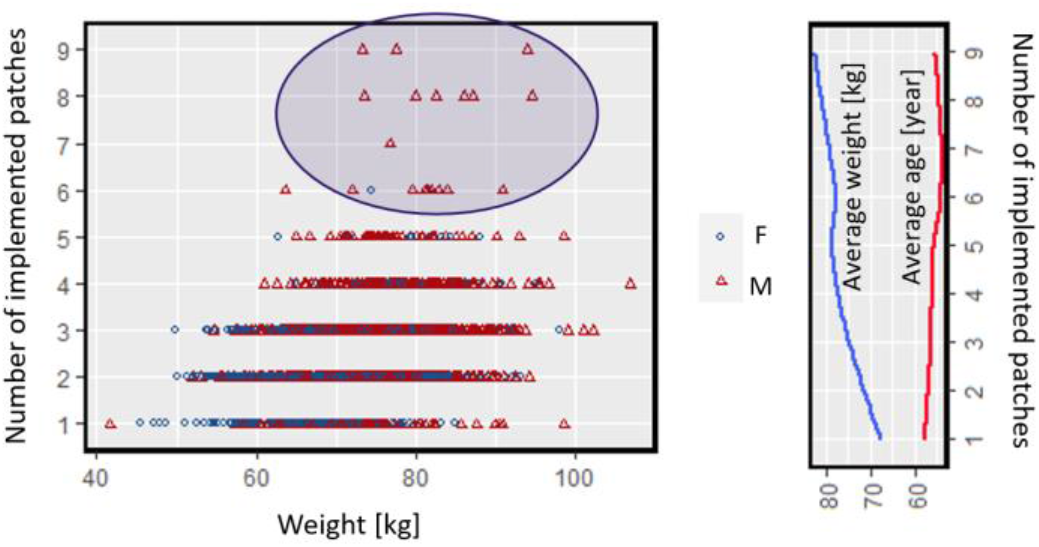
a. number of implemented patches for each virtual patient in 72 hours during digital-twin-assisted therapy based on weight, b. average weight and age of the patients, based on the number of patches they used in 72 hours during digital-twin-assisted therapy.

In the next step, we analyzed the patients that needed to change the patch more frequently. We separated the part of the population that they need more than one patch per day. We compare the patient physiology distribution between the population and this subgroup. The result in Figure 10 shows that the two groups’ age distribution is not significantly different (p_value =0.45). However, for weight distribution, there is a shift in the mean of the distribution (by +6 kg), and with a p_value of 2.2 * 10^−5^, these two distributions are different. For height distribution, the mean of the distribution is shifted by 3 cm, and the two distributions are significantly different (p_value=0.004). The ratio of females to males in the population is 1.02; however, in the subgroup, this ratio is 0.29. Based on this result, patients of the male gender and with higher body mass need to change the fentanyl patch more frequently to reach the target pain relief.

**Figure 10.**
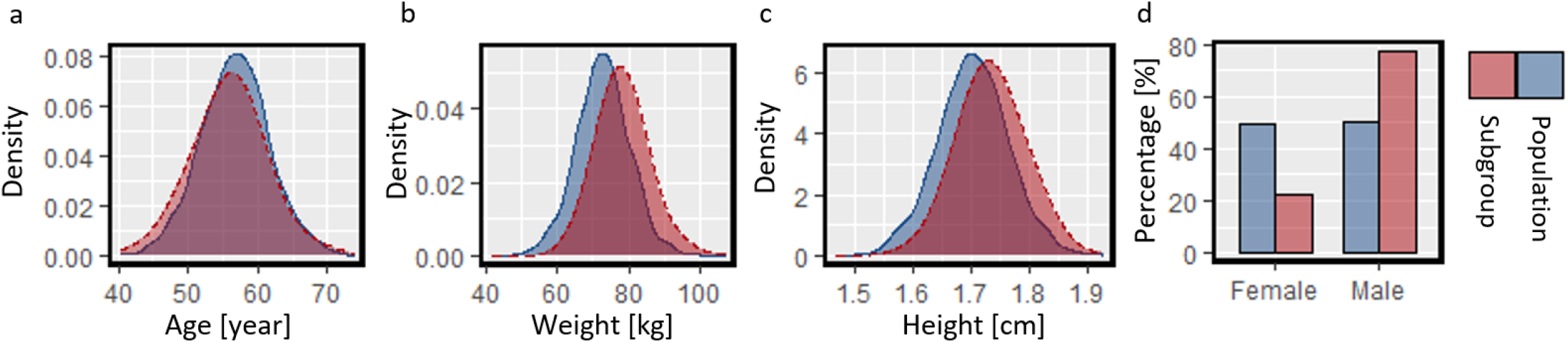
a. age distribution, b. weight distribution, c. height distribution, d. gender percentage for the population and the subgroup of patients needing to use more than one patch a day.

## 4 OUTLOOK

This study analyzed the patient characteristics that affect fentanyl transdermal therapy. These characteristics were age, weight, gender, and height. Based on patient physiological features and published experimental studies, model parameters for drug uptake, pharmacokinetics, and pharmacodynamics model were calculated. In the next step, the tailored digital twin for each patient proposed a modification of the therapy based on the pain intensity. Digital-twin-assisted therapy successfully reduced the pain intensity and the variation in therapy outcomes for virtual patients. However, there are approaches to increase the compatibility to the real-world and higher efficiency and safety in the therapy. These approaches are mentioned below:

- In order to reduce the error in the calculation and prediction of the digital twin, the model parameters should be calculated based on the result of fentanyl therapy on the patient.
- Liver enzyme activity and kidney filtration rate greatly impact the plasma concentration level as they determine the elimination and metabolism rate of fentanyl [47]. These physiological features must be considered in tailoring the digital twin for the patient.
- In this study, only pain intensity was considered the main factor in modifying the therapy. However, in order to reach a safer therapy, it is important to consider adverse effects such as hypoventilation in decision-making.
- Besides respiratory depression, fentanyl transdermal therapy can cause other side effects such as nausea, skin dryness, muscle spasms, confusion, and anxiety [43]. Taking to account these side effects and trying to avoid them while having efficient pain relief will lead to more tolerable therapy for the patient.

## 5 CONCLUSIONS

In this study, we tailored a physics-based digital twin of transdermal fentanyl therapy for 3000 virtual patients. By implementing a set of sample data and Markov chain Monte Carlo (MCMC), these virtual patients that were different from each other based on age, gender, weight, and height were developed. The model parameters for each patient were calculated based on these physiological features and applying interindividual variability. Conventional SmPC-based therapy of fentanyl transdermal patch, which is implementing fentanyl patch and replacing it every 72 hours, was studied. Based on the outcome of conventional therapy, the maximum fentanyl concentration varied in the range of 1.3 -3.2 ng ml^-1^. Additionally, the minimum VAS pain score varied in the range of 0-5, and the minimum minute ventilation rate was in the range of 2.5-10 L min^-1^ for the virtual patients. This result shows that the outcome of therapy has a high variation between patients as some patients receive excessive amounts of fentanyl while some of the virtual patients could not reach the target pain relief.

In order to reach the target pain relief for each patient, we designed a new therapy by digital twin. In this digital-twin-assisted therapy, the patient’s pain intensity will be checked every 8 hours, and if it is higher than the target, the patient implements a new patch of the same size. As a result of this modification, the average pain intensity by 16% and the standard deviation by 54% decreased. Based on the result, the digital twin successfully kept the pain intensity for 98.8% of the patients below 3 for more than half of the treatment duration. However, this number is only 57.1% for conventional therapy. Digital-twin-assisted therapy increased the median time without pain (VAS pain score under 3) by 23 hours in 3 days and reduced its interquartile range (IQR) by 70%. Based on this result, digital-twin-assisted therapy reduces the overall pain intensity for the whole virtual population and reduces the variation of the outcome of therapy between the patients. Therefore, the tailored digital twin can predict the patients’ needs and conditions, propose a therapy, monitor the endpoint of therapy, and update when the patient needs it. In this way, all patients suitable for fentanyl transdermal therapy can receive suitable pain relief.

## Data Availability

The human data is from the published study of Zech et al. in 1992: https://doi.org/10.1016/0304-3959(92)90034-9
The simulated data of the study are available upon reasonable request to the authors

## Acknowledgments

This work was financially supported by the Novartis Research Foundation (grant “Virtual twinning for intelligent, personalized transdermal drug delivery”) and OPO Foundation (grant “ Digital human avatars help tailor transdermal pain management (TREATME)”). The funders were not involved in the study design, collection, analysis, and interpretation of data, the writing of this article, or the decision to submit it for publication.

